# Initial Model for the Impact of Social Distancing on CoVID-19 Spread

**DOI:** 10.1101/2020.05.04.20091207

**Authors:** Genghmun Eng

**Affiliations:** PhD Physics 1978, University of Illinois at Urbana-Champaign

## Abstract

The initial stages of the CoVID-19 coronavirus pandemic all around the world exhibit a nearly exponential rise in the number of infections with time. Planners, governments, and agencies are scrambling to figure out “*How much? How bad?*” and how to effectively treat the potentially large numbers of simultaneously sick people. Modeling the CoVID-19 pandemic using an exponential rise implicitly assumes a nearly unlimited population of uninfected persons (*“dilute pandemic”*). Once a significant fraction of the population is infected (*“saturated pandemic”*), an exponential growth no longer applies. A new model is developed here, which modifies the standard exponential growth function to account for factors such as *Social Distancing*. A *Social Mitigation Parameter [SMP] α_S_* is introduced to account for these types of society-wide changes, which can modify the standard exponential growth function, as follows:

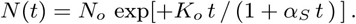

The *doubling-time t_dbl_* = (In 2)/*K_o_* can also be used to substitute for *K_o_*, giving a {*t*_dbl_, *α_S_*} parameter pair for comparing to actual CoVID-19 data. This model shows how the number of CoVID-19 infections can self-limit before reaching a *saturated pandemic* level. It also provides estimates for: (a) the timing of the *pandemic peak*, (b) the maximum number of new daily cases that would be expected, and (c) the expected total number of CoVID-19 cases. This model shows fairly good agreement with the presently available CoVID-19 pandemic data for several individual States, and the for the USA as a whole (*6 Figures*), as well as for various countries around the World (*9 Figures*). An augmented model with two *Mitigation Parameters, α_S_* and *β_S_*, is also developed, which can give better agreement with the daily new CoVID-19 data. Data-to-model comparisons also indicate that using *α_S_* by itself likely provides a worst-case estimate, while using both *α_S_* and *β_S_* likely provides a best-case estimate for the CoVID-19 spread.

## 1 Introduction

The Coronavirus 2019 disease (CoVID-19), caused by the SARS-CoV-2 (Severe Acute Respiratory Syndrome Coronavirus 2) pathogen, is now a world-wide pandemic. In many localities, the number of cases *N*(*t*) was found to have an initial period of nearly exponential growth:

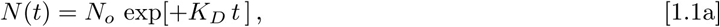

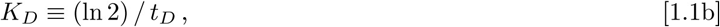

aside of the first few cases, which may be untraceable. In Eqs. [1.1a]-[1.1b], *N_o_* is the initial number of infections at the *t* = 0 start of data tracking, *K_D_* is an exponential growth factor, and *t_D_* is the *doubling-time*. Each locality can have its own {*N_o_*, *t_D_*, *t* = 0} values, and *K_D_* and *t_D_* should be nearly constant during this initial period of CoVID-19 spread.

Standard epidemiology identifies the number of people *N*_G_ a known infected individual had recent contact with. Contacts of that *N*_G_ group are tracked next, followed by additional tracking stages. This process sets the *K*_D_ value.

Society-wide *Mitigation Measures* such as: (a) *Social Distancing*, (b) wearing face masks in public, (c) prohibiting large gatherings, (d) implementing large-scale population testing, (e) disinfecting high-touch surfaces in public areas, (f) enhanced cleaning of items brought into homes, and (g) minimizing human contact with likely virus-containing materials and matter; all can help reduce *N*(*t*) growth. These *Mitigation Measures* can modify the Eq. [1.1a] epidemiology model by causing the local *t_D_* values to lengthen.

In order to model these *Mitigation Measures, t_D_* and *K_D_* become explicit functions of time, *t_D_* (*t*) and *K_D_* (*t*). Using a linear function for *t_D_* (*t*) lengthening is one of the simplest time-varying extensions. A linear function of time also corresponds to the first term of a *Taylors’ Series* expansion of some more general *t_D_* (*t*) analytic function, giving this epidemiology extension:

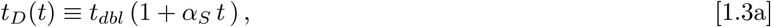

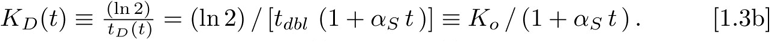

The *t* = 0 initial values for *t_D_* (*t*) and *K_D_* (*t*) become the new constants *t_dbl_* and *t_D_*, which characterize the initial exponential growth phase. The *α_s_* coefficient in Eq. [1.3a] is a new *Social Mitigation Parameter [SMP]* that helps quantify the effectiveness of the society-wide *Mitigation Measures* as a whole.

The *α_s_* value expresses how well non-infected people manage to avoid the virus contagion. As a lumped parameter, it likely reflects an average value over many processes, known and unknown, which comprise mitigation, to supplement the contact-to-contact tracking that initially sets *t_dbl_* or *K_o_*.

Substituting Eq. [1.3b] into Eq. [1.1a] gives:

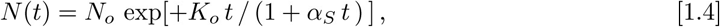

as one of the simplest models for CoVID-19 spread. A pure exponential growth (or decay) has no *memory*, while Eq. [1.4], for *α_S_* > 0, has a *memory*. The *t* = 0 start time of first mitigation changes the future history. To include *t*< 0 requires replacing Eq. [1.3a] by *t_D_*(*t*) = *t_dbl_* (1 + max[0, *α_S_ t*]), which has a corner at *t* = 0 that preserves the *memory* of when mitigation first started.

## 2 Model Features

The Eq. [1.1a] exponential growth pandemic model implicitly assumes a large uninfected population allows the disease to easily spread (“*dilute pandemic”*). When almost everybody is infected (“*saturated pandemic”*), exponential growth shuts off, and Eq. [1.1a] no longer applies.

On 3/10/2020, *German Chancellor Angela Merkel***^1^** noted that she *“estimates that 60% to 70% of the German population will contract the coronavirus*”, indicating that *saturated pandemic* models are being considered as a worst-case.

Even that worst-case condition assumes: (i) recovered coronavirus patients are no longer infectious, and (ii) surviving a CoVID-19 infection confers absolute immunity to re-infection. Recently, South Korea**^2^** found 91 cases of clinically recovered patients later testing as CoVID-19 positive. They may also shed viable coronaviruses in their phlegm and fecal matter**^3^**, furthering disease spread. Although these effects are not modeled here, those additional transmission modes could turn a 60%-70% hope into a 99+% consequence.

These factors show why CoVID-19 modeling beyond Eq. [1.1] is needed, especially to see if society-wide *Mitigation Measures* can naturally halt disease spread, without necessitating a *saturated pandemic* condition. We show next that Eq. [1.4] allows for this pandemic shut-off, even in the *dilute pandemic* case. Since both *K_o_* and *α_S_* in Eq. [1.4] have the same units, their ratio is dimensionless. The long-term limit of Eq. [1.4] gives:

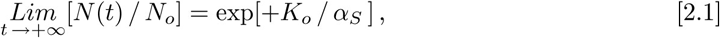

setting an average value for the total number of all follow-on infections arising from a single individual. Since it depends only on the ratio of the original pandemic growth factor *K_o_* to the *α_S_ SMP*, this model shows the impact of accounting for a broader environment beyond individual contact tracking.

The early spread of CoVID-19 cases outside of China**^4^**, and the early USA CoVID-19 data**^5^** both had nearly exponential rises, as shown in **Figure 1**. A purely exponential rise gives a straight line on a *log-plot* {*log*(*# of cases*) *vs linear time*}. The initial *doubling-time* for the USA was *t_dbl_* ≈ 2.02 *days*, giving *K_o_* ≈ 0.343 */ day* using Eqs. [1.1a]-[1.1b]. This initial CoVID-19 data was prior to any significant *Mitigation Measures* being implemented.

**Figure 1:**
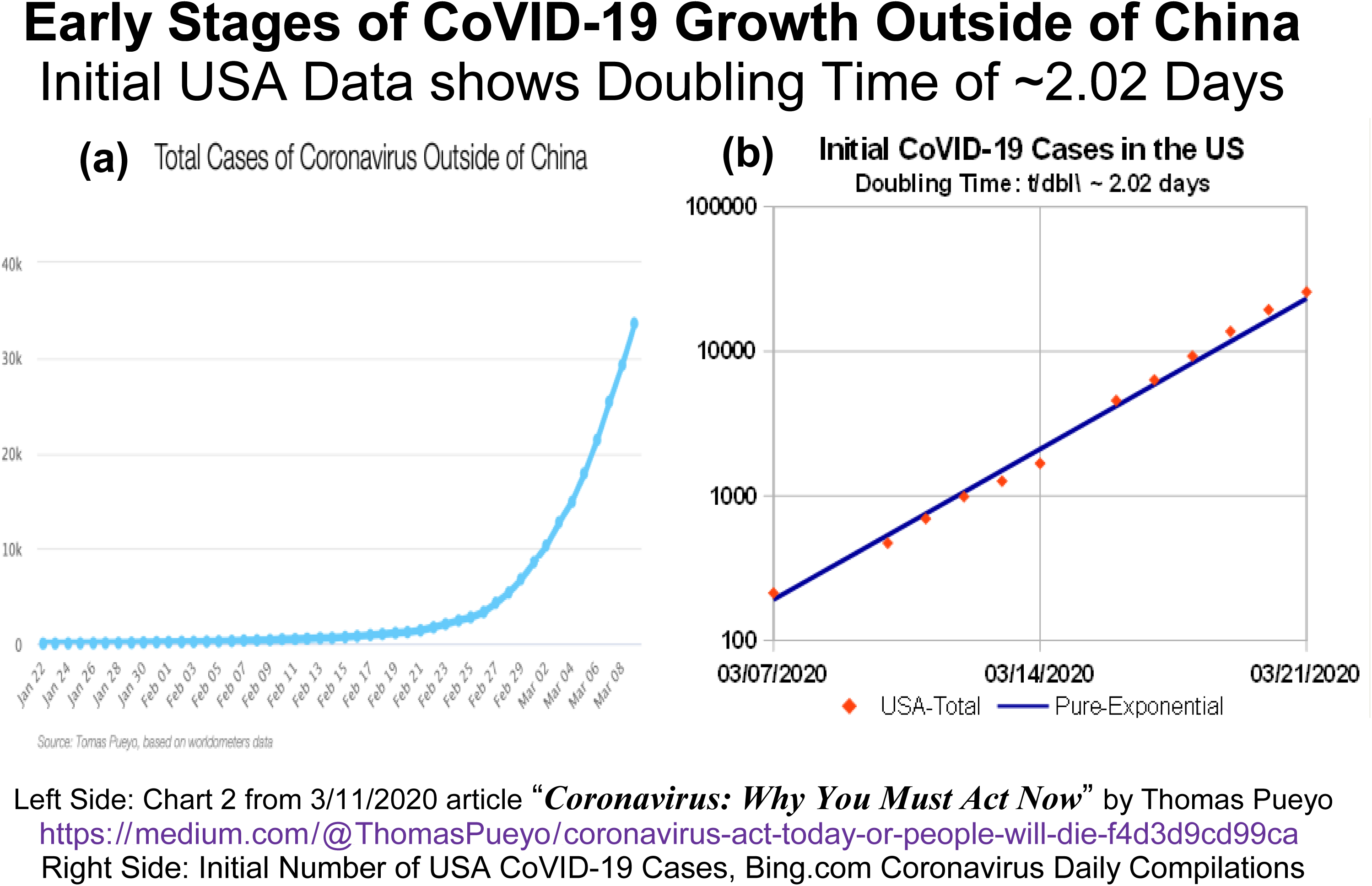
Early CoVID-19 Cases: (a) Outside of China, and (b) Just in the US. Both graphs show nearly exponential growth.

On March 19, 2020, *Governor Gavin Newsom* of California ordered a CoVID-19 “stay-at-home” lockdown of virtually all of California’s ^~^40 million residents. Similar statewide CoVID-19 lockdowns were ordered by the Governors of Illinois, New York, Indiana, Michigan, Ohio, Washington, West Virginia and Wisconsin.

The slowing of CoVID-19 spread by implementing large-scale societal *Mitigation Measures* can be fairly rapid, as illustrated by the USA CoVID-19 data of **Figure 2**, which covers March 2020.

**Figure 2:**
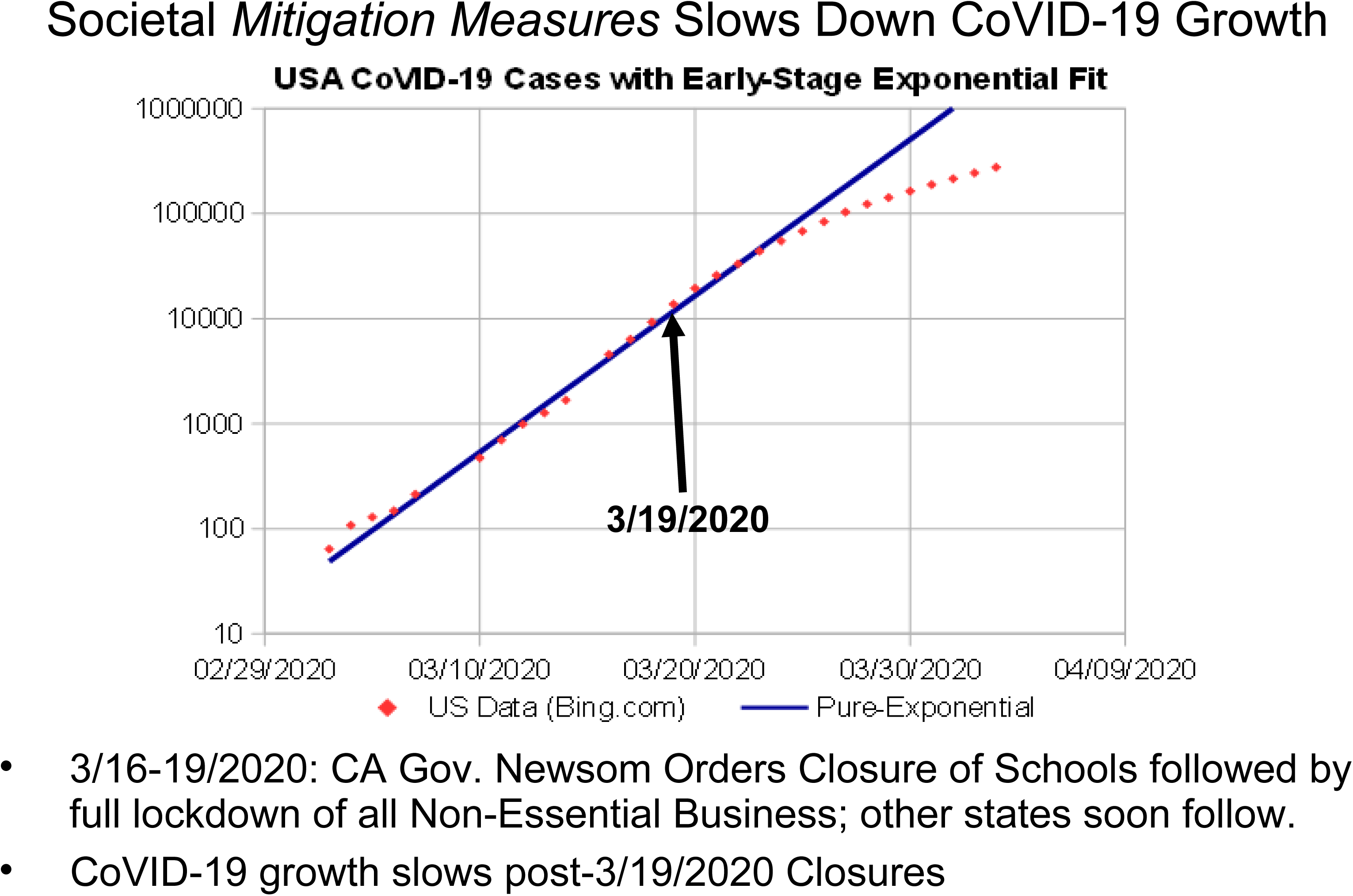
USA CoVID-19 data, pre-vs-post mid-March 2020. Multi-State *Mitigation Measures* slowed growth, transitioning from straight-line to downward curvature.

The impact of these multi-state *Mitigation Measures* is evident in **Figure 2** as a sudden transition on the *log-plot* from a straight-line to having downward curvature, which the as *Social Mitigation Parameter (SMP)* aims to quantify. The local slope in **Figure 2** also decreases right after the onset of *Mitigation Measures*, indicating further slowing of CoVID-19 spread.

A well-documented South Korean coronavirus cluster can also be used to help estimate the expected size of the *α_S_ SMP*. That CoVID-19 cluster determined that a single infected person at the *Shinjeongji Church* caused infection of about 4, 482 people**^4^** within the 47-day time interval between January 20, 2020 and March 8, 2020. Those data provide this *α_S_* estimate:

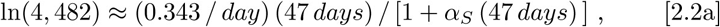

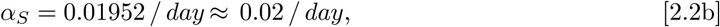

as indicative of minimal mitigation. If additional deliberate mitigation measures doubled *α_S_* to *α_S_* ≈ (0.04 / day), Eq. [2.1] would give:

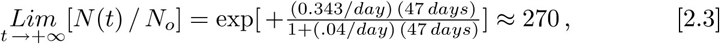

for the number of infections per person, a 16.6 *X* reduction from 4, 482.

Since *N*(*t*) in Eq. [1.4] represents a total number of cases, it is similar to a *cumulative distribution function (cdf)*, which is used in reliability and also has time as its fundamental variable. The derivative of Eq. [1.4], *dN*(*t*) / *dt*, is analogous to an unnormalized *probability density function (pdf)*, which can be used to predict a *pandemic peak*:

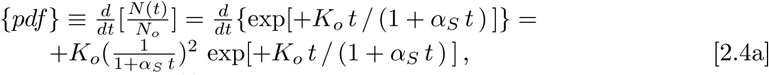

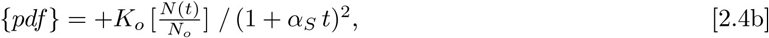

The time *t_P_* of the *pandemic peak* is set by:

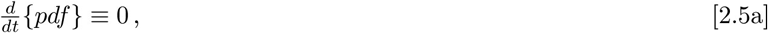

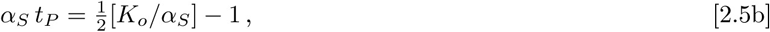

where Eq. [2.5b] simplification arises from the Eq. [2.5a] constraint. Substituting *K_o_* = 0.343 / day and *α_S_* = 0.02 / day from Eq. [2.2b] into Eq. [2.5b] gives *t_P_* ≈ 379 days. Increasing mitigation to *α_S_* = 0.04 / day, keeping the same *K_o_* = 0.343 / day, now gives *t_P_* ≈ 82 days, which is a ^~^4.6 *X* reduction in the *pandemic peak* timing for doubling (2*X*) the mitigation effect from its original baseline value. These examples highlight the tremendous impact that even a small amount of enhanced social mitigation can have.

While the *α_S_* =0 limit of Eq. [2.4b] recovers the Eq. [1.1a] standard exponential growth, both the {*pdf*} and [*N*(*t*)/*N_o_*] growth are then unbounded. However, even a small *α_S_* > 0 value in Eq. [2.4b] will have an enormous impact on the predicted long-time behavior. Since Eq. [2.1] showed that [*N*(*t*)/*N_o_*] now approaches a finite value for all *α_S_* > 0, this new {*pdf*} asymptotic limit:

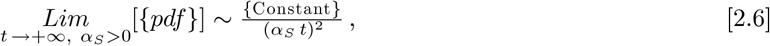

also results. The {*pdf*} that arises from this model all have an initial exponential rise, coupled with the Eq. [2.6] “*long tail*” at large times, which means that new CoVID-19 cases may arise for a long time, even if significant *Mitigation Measures* are in place.

The Eq. [2.6] {*pdf*} prediction also differs substantially from the widely-used *University of Washington IHME (Institute for Health Metrics and Evaluation*) projections, which use symmetric Gaussians for both the {*pdf*} rise and fall**^6^**. Thus, these methods provide an alternative risk-bound for evaluating potential CoVID-19 worst-case scenarios.

## 3 Determining {*t_dbl_*, *α_S_*} from CoVID-19 Data

Explicit numerical values for {*t_dbl_, α_S_* } parameters were determined from the CoVID-19 data as follows. Rewriting Eq. [1.4] as:

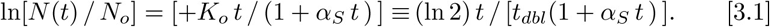

allowed data fitting to be done on a *Y – vs – X log-plot*, using *Y* = ln[*N*(*t*) / *N_o_*] as the ordinate and *X* ↔ *t* as the abscissa, to calculate and minimize the *root-mean-square (rms) error*.

The *t* = 0 point in Eq. [3.1] sets *N_o_*. To best model *Mitigation Measures*, this point was usually chosen at the start of a downward curvature on a *log-plot*, so that *N*(*t* = 0) = *N_I_*, where the *N_I_* is now the first data point in the analysis. The prior *t <* 0 regime can often have a nearly pure exponential growth, as in **Figure 2**, and those regions should not be part of *rms-error* minimization for evaluating *Mitigation Measures*.

The *N_F_* final data point, measured at the most recent *t* = *t_F_* time:

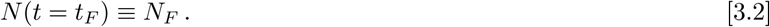

was also fixed for each dataset, so that only {*t_dbl_, α_S_*} value pairs that meet both *N(t* = 0) = *N_I_* and Eq. [3.2] were used.

In practice, an *α_S_* was chosen first. The *Excel™_Tools_Goal-Seek* function was used to adjust *t_dbl_* to obey Eq. [3.2], setting the *rms-error* between the dataset and Eq. [3.1], with the final {*t_dbl_, α_S_*} having the least *rms-error*.

In the following figures, all CoVID-19 raw data came from the publicly available Microsoft™ “ *COVID-19 Tracker*” site**^7^**. When no updates were available, that site repeated the prior day data, whereas we used the geometric mean of the day-prior and day-after data for interpolation.

## 4 Effects of Varying the Initial Zero-Time Point

Starting with:

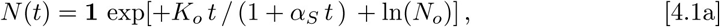

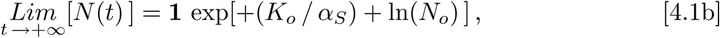

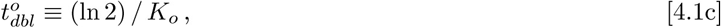

using a shifted time-scale normalization point is examined next:

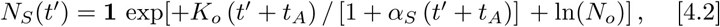

This *N_S_*(*t*ʹ) function should closely match Eq. [4.1a], with a shifted time axis: *t* = *t*ʹ + *t_A_*, but the best fit parameter numerical values change. Since:

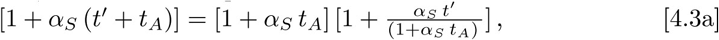

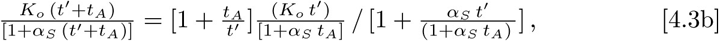

then defining:

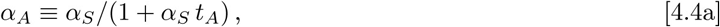

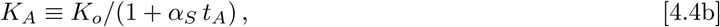

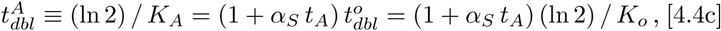

it gives:

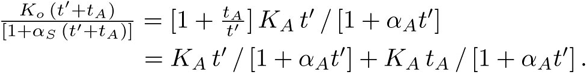

These equations highlight the net effect of time-shifting. For *t_A_* > 0, when *t*ʹ begins after *Mitigation Measures* have started, the shifted-time-axis results in a larger calculated *doubling-time* and a smaller *SMP α_A_*-value. For *t_A_* < 0, when *t*ʹ may include *Mitigation Measures* already in place at the Eqs. [4.1a]-[4.1c] *t* = 0 point, this shifted-time-axis results in a smaller calculated *doubling-time* and a larger *SMP α_A_*-value.

Finally, for small *t*ʹ, where 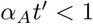, using Eqs. [4.1b] and [4.5] gives:

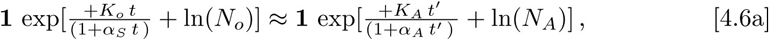

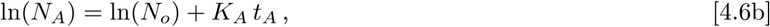

which shows that the *t*ʹ = 0 new initial state should have an *N_A_* starting value obeying *N_A_* > *N_o_* for *t_A_* > 0, and *N_A_* < *N_o_* for *t_A_* < 0. However, whether {*N_o_*, *K_o_*, *α_S_*}, or an alternative {*N_A_*, *K_A_*, *α_A_*}, are used to parameterize a given data set, the net overall function fit and predictions, as a function of calendar date, should remain fairly self-consistent, even when some ambiguity exists as to when *Mitigation Measures* first were noticeably effective.

## 5 USA and Selected States Model Results

The model predictions for CoVID-19 spread in the USA is shown in **Figure 3**. This analysis only included data after mid-March 2020, when several State Governors first instituted mandatory *Mitigation Measures*. Results give an *SMP* estimate of *α_S_* ≈ 0.5945/*day*, a USA initial *doubling-time* of 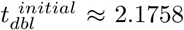 *days*, which lengthens to 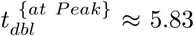 *days* at the projected *pandemic peak* of ^~^4/19/2020. The predicted total number of CoVID-19 cases is ^~^5, 464, 000, giving a projected ^~^1.67% final infection rate, if the present level of *Mitigation Measures* or their equivalent, are continued.

**Figure 3:**
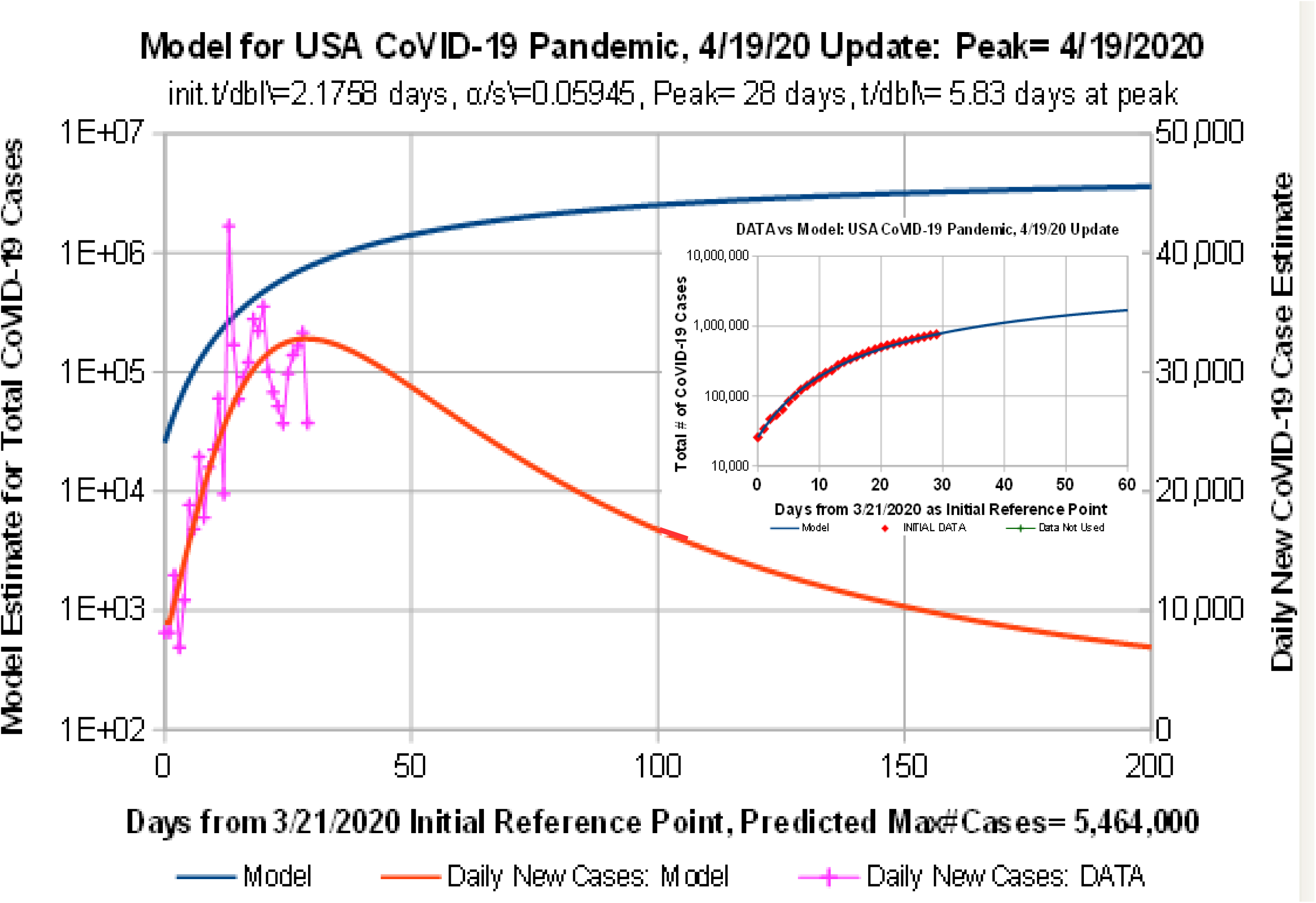
USA Model Predictions. To allow better mitigation predictions, only data after mid-March 2020 was included, when several Governors instituted mandatory lockdowns.

These predictions assume no *“second wave”* of infection or re-infection. They also do not include the effect of additional *Mitigation Measures*, which could further increase the {*t_dbl_*, *α_S_* } values, and significantly reduce the projected final number of CoVID-19 cases.

**Figure 4** shows model predictions for CoVID-19 evolution in California. Only data after 3/21/2020 was included in the analysis, after California Governor Gavin Newsom instituted mandatory *Mitigation Measures*. It gives an *SMP* estimate of *α_s_* ≈ 0.03546/*day*, with an initial *doubling-time* of 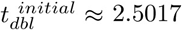 *days*, which lengthens to 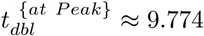 *days* at the projected *pandemic peak* of ^~^6/07/2020. The predicted total number of CoVID-19 cases is ^~^1,123, 700, giving a projected ^~^2.813% final infection rate, at the present level of *Mitigation Measures*.

**Figure 4:**
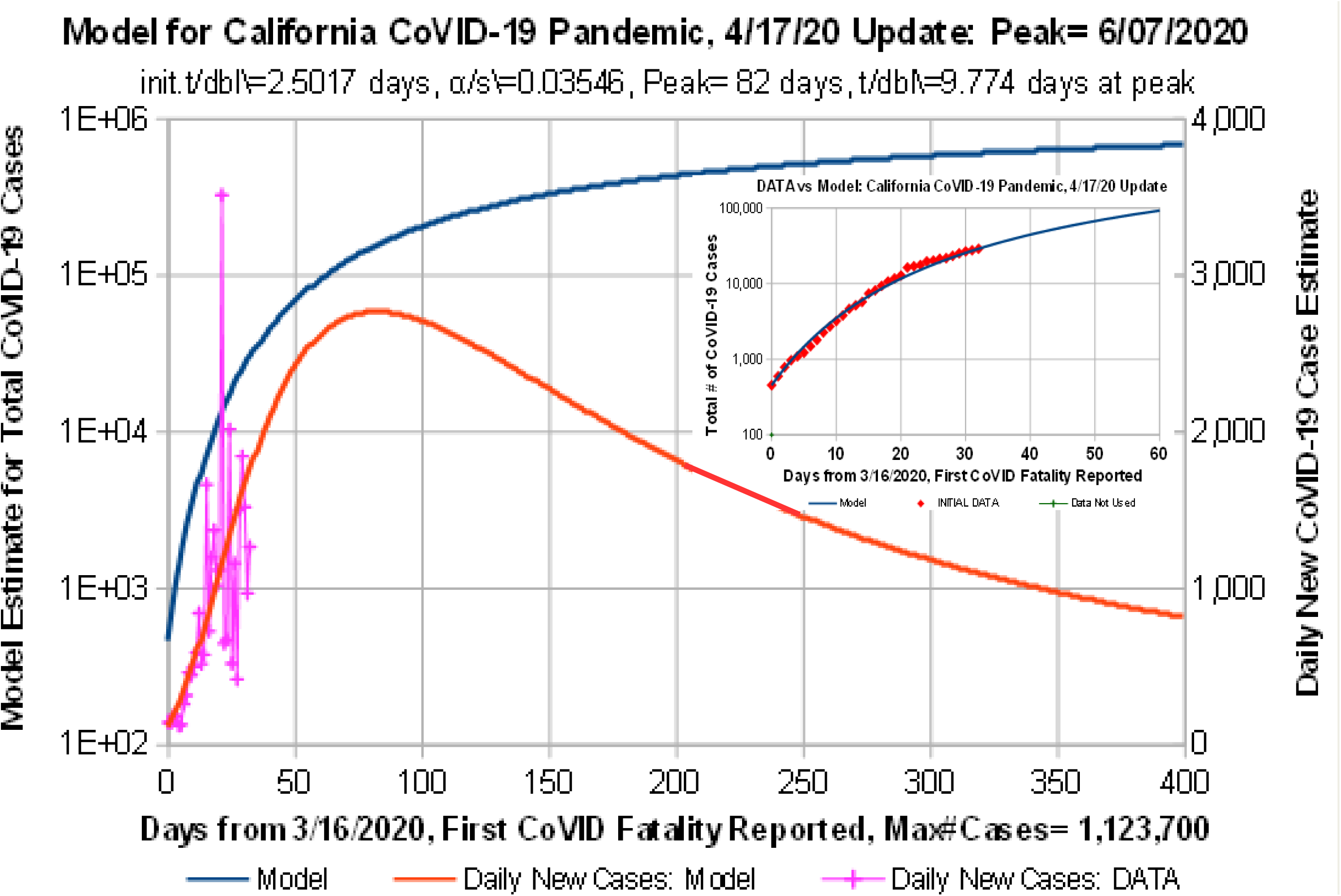
Predicted California CoVID-19 results. After Gov. Gavin Newsom instituted widespread *Mitigation Measures*, projections showed significant improvement.

**Figure 5** shows model predictions for CoVID-19 evolution in New York. A relatively high *SMP* estimate of *α_s_* ≈ 0.1031/*day* was found, coupled with a relatively short initial *doubling-time* of 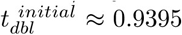 *days*, which creates a high narrow spike in daily new cases. The present model projects a New York *pandemic peak* around 4/10/2020, with an estimated at-peak *doubling-time* of 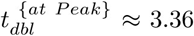 *days*. The predicted total number of cases at ^~^1, 218, 000, giving a projected ^~^6.072% final infection rate.

**Figure 5:**
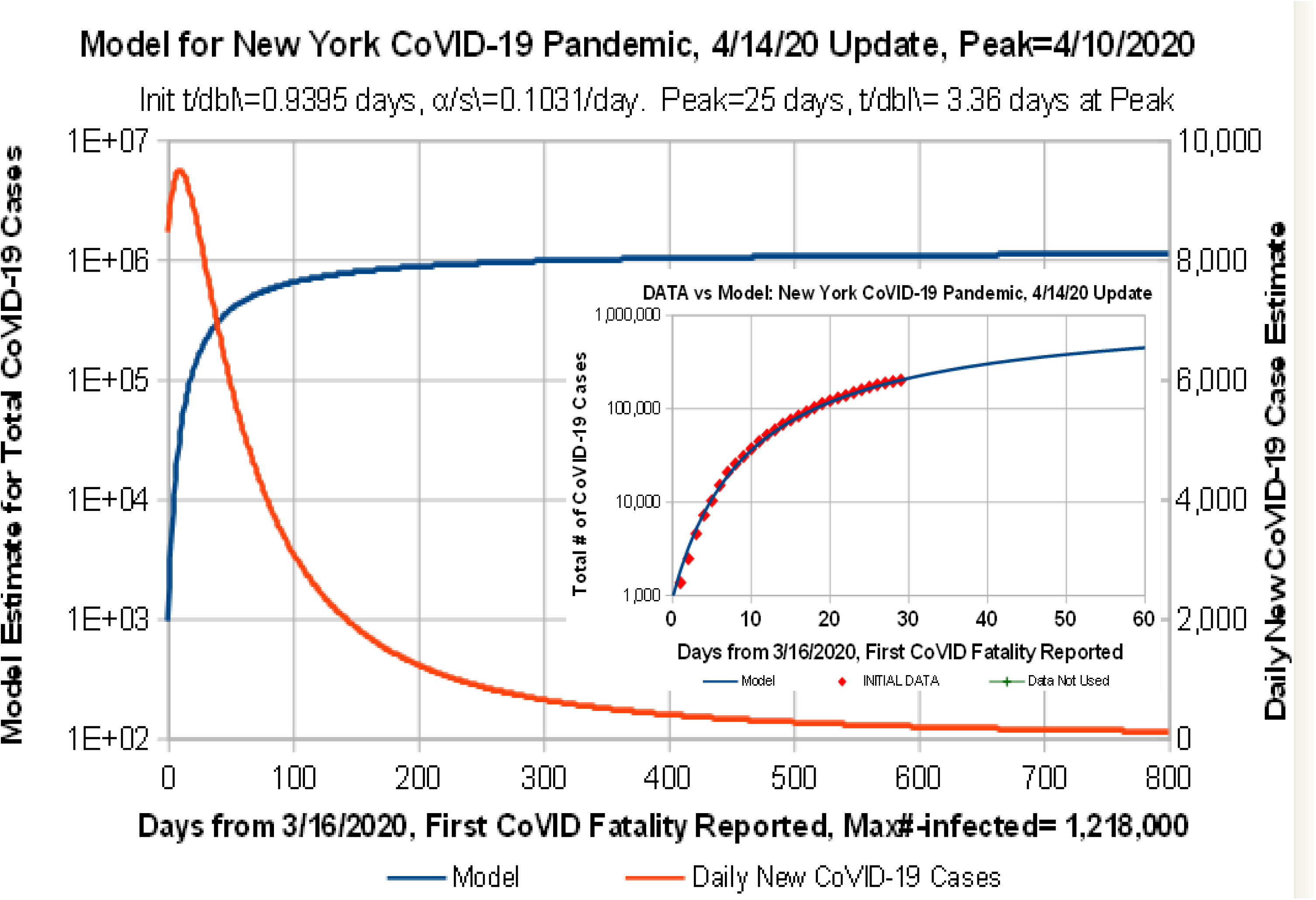
Predicted New York CoVID-19 results. A relatively high *Mitigation Measure* level and a short intrinsic doubling time creates a narrow spike in daily new cases.

**Figure 6** shows model predictions for CoVID-19 evolution in Washington State. An initial *doubling-time* of 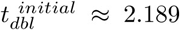 *days* and an *SMP* value of *α_S_* ≈ 0.0687/*day* were found, with a projected *pandemic peak* around 6/04/2020. The relatively low number of cases at the *Mitigation Measures* start helps to give a predicted total number of cases of ^~^557, 600, corresponding to a ^~^7.15% final infection rate.

**Figure 6:**
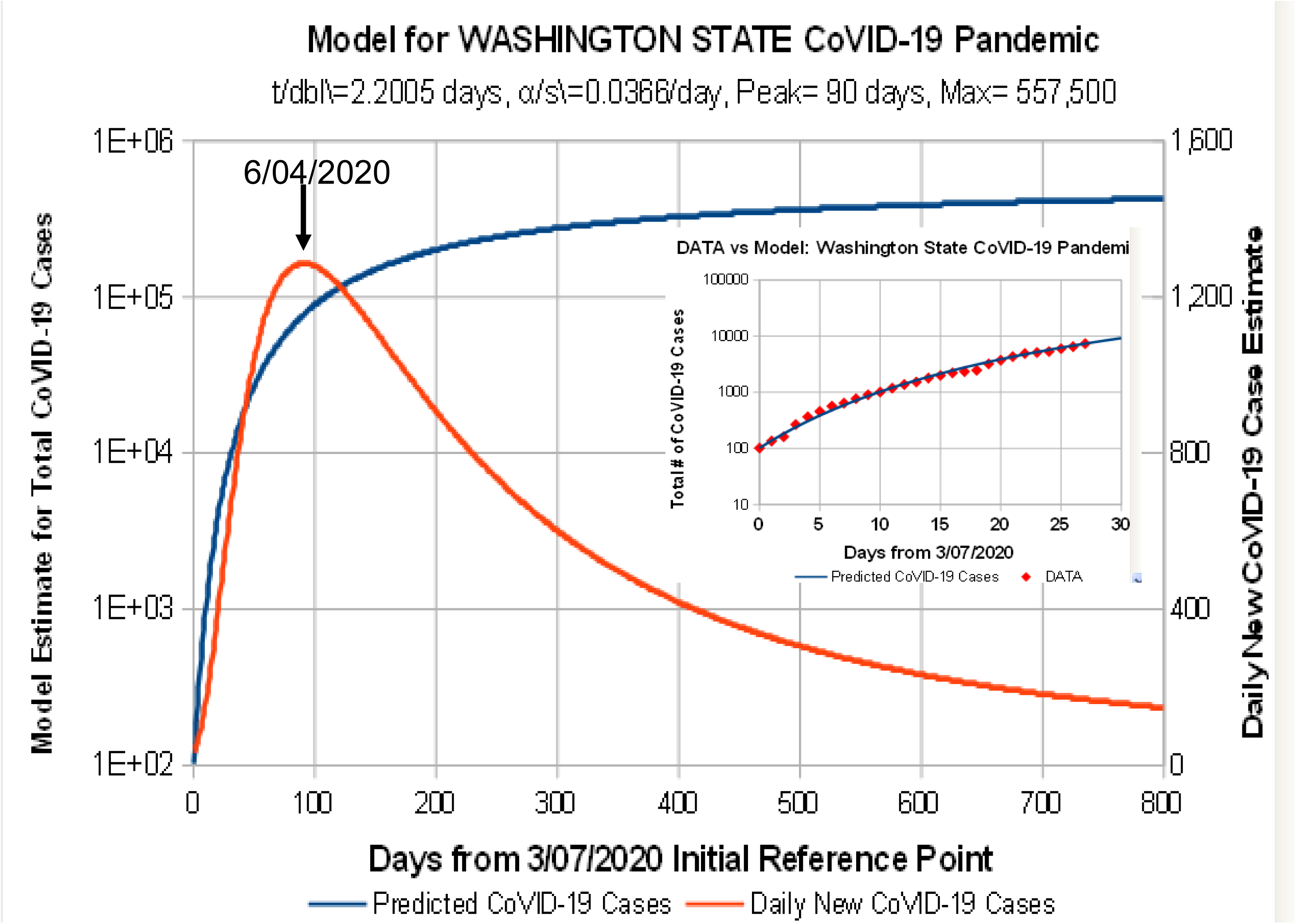
Predicted Washington State CoVID-19 results. The relatively low number of cases at *Mitigation Measure* start helps to give a relatively low final number of cases.

**Figure 7** shows model predictions for CoVID-19 evolution in Illinois. An initial *doubling-time* of 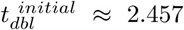 *days* and moderate *SMP* value of *α_S_* ≈ 0.0373*/day* combine to give a projected *pandemic peak* around 6/04/2020, similar to Washington State, but having a higher predicted total number of cases at ^~^1, 277, 000, and a projected ^~^11.47% final infection rate.

**Figure 7:**
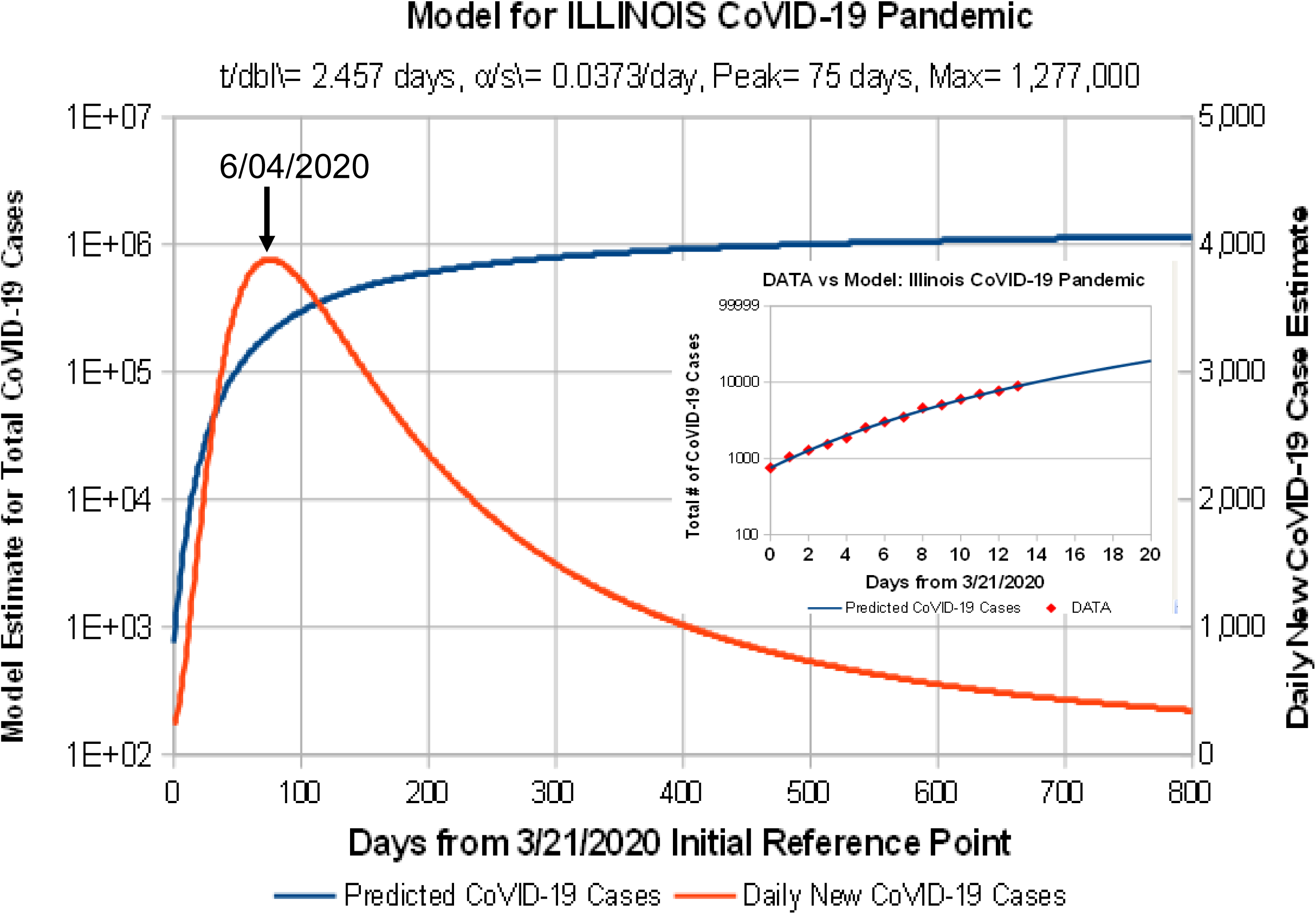
Predicted Illinois CoVID-19 results. The slow *doubling-time* and moderate amount of *Mitigation Measures* gives a slow increase to the predicted CoVID-19 peak.

**Figure 8** shows model predictions for CoVID-19 evolution in Florida. Many Florida counties instituted their own Mitigation Measures prior to a state-wide lockdown, slowing CoVID-19 growth. A somewhat high *SMP* value of *α_S_* ≈ 0.0526/*day*, and an initial *doubling-time* of 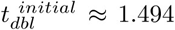*days* results. A *pandemic peak* is estimated at around 5/20/2020, with a predicted total number of cases at ^~^1, 090, 000, and a projected ^~^4.96% final infection rate.

**Figure 8:**
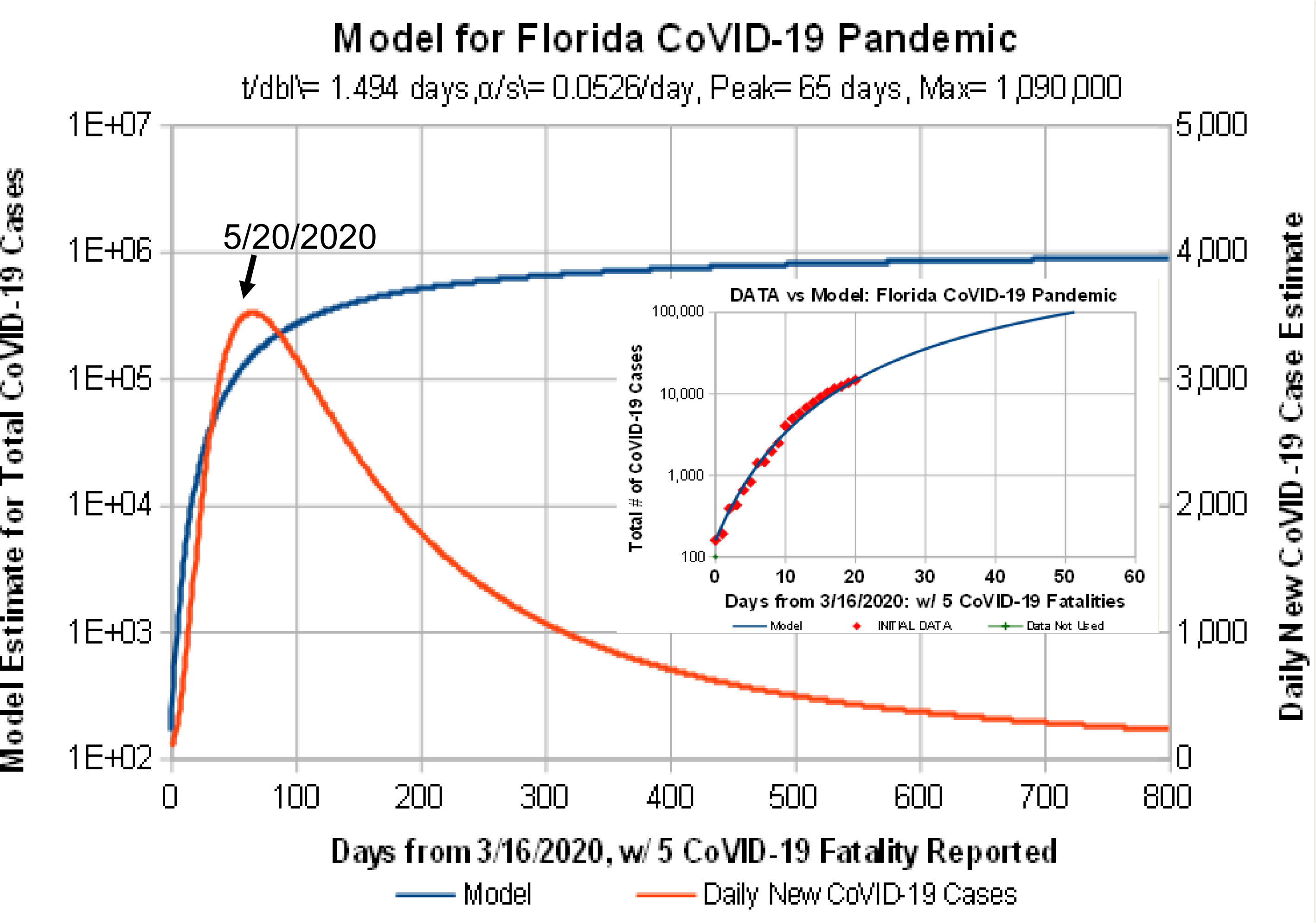
Predicted Florida CoVID-19 results. Many Florida counties instituted their own Mitigation Measures prior to a state-wide lockdown, substantially slowing CoVID-19 growth.

## 6 World and Selected Countries Model Results

**Figure 9** shows model predictions of CoVID-19 evolution for the whole World. The present-day *doubling-time* value of 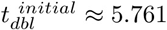 *days* likely represents a combination of small urban, large urban, and rural area results. However, the calculated low *SMP* estimate of *α_S_* ≈ 0.01712/*day* shows that nearly ^~^4.43% of the World’s population could be at risk for eventual CoVID-19 infection. At these present levels, the projected *pandemic peak* is around 8/15/2020, with potentially hundreds of millions of people being infected.

**Figure 9:**
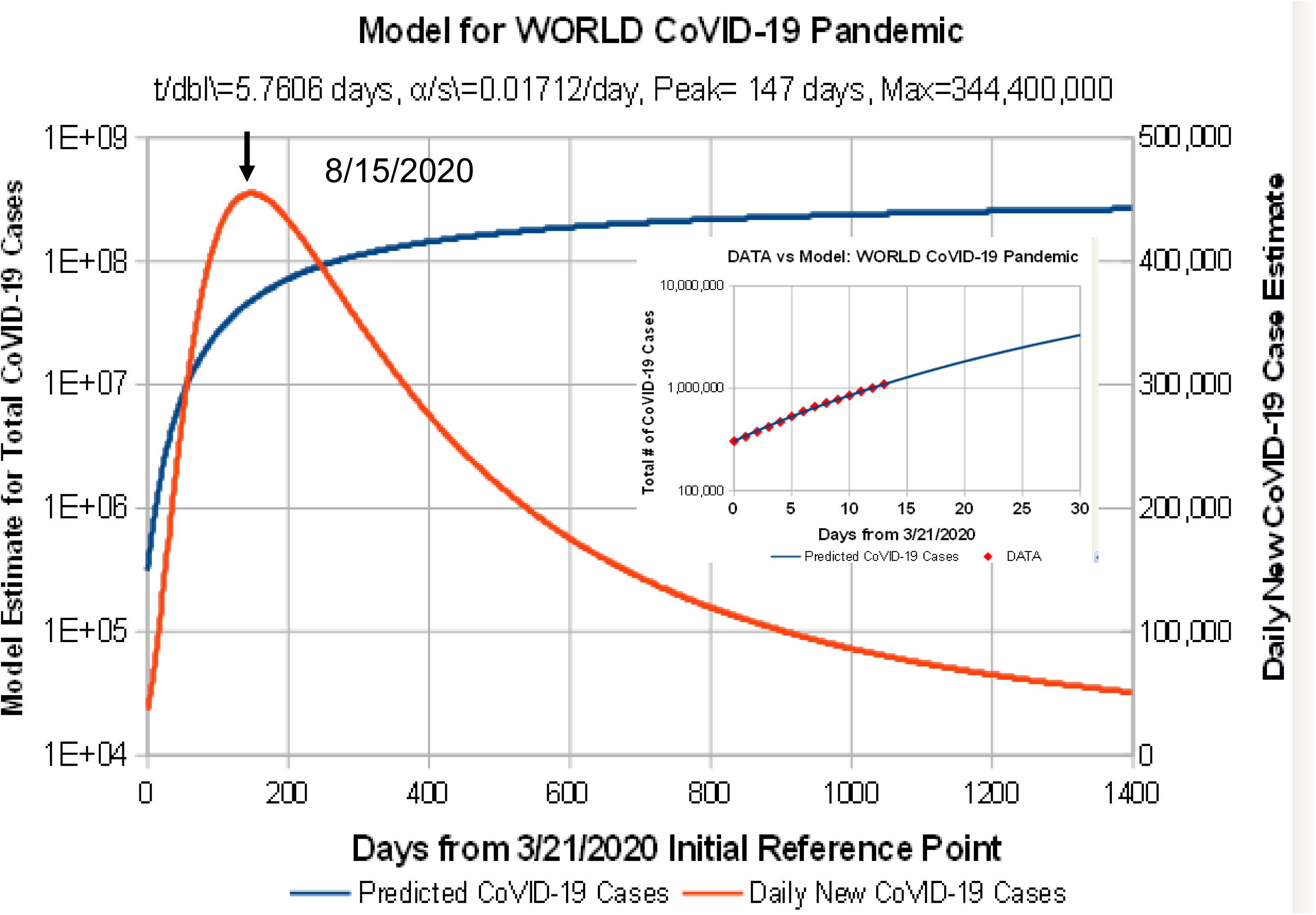
Model predictions for the WORLD, showing present-day low level of mitigation.

**Figure 10** shows model predictions for CoVID-19 evolution in China, covering their *“first wave*” of early exposure and early mitigation. Data were included that was prior to a “*New Reporting Method*” being used, which started off with one sudden data jump, and nearly level CoVID-19 follow-on results. The present model predicts what number of cases could have resulted, had the reporting method not changed. Draconian *Mitigation Measures* helped to contain the pandemic to *Hubei Province* and *Wuhan*. These projections show that those *Mitigation Measures* have impressively contained CoVID-19 spread.

**Figure 10:**
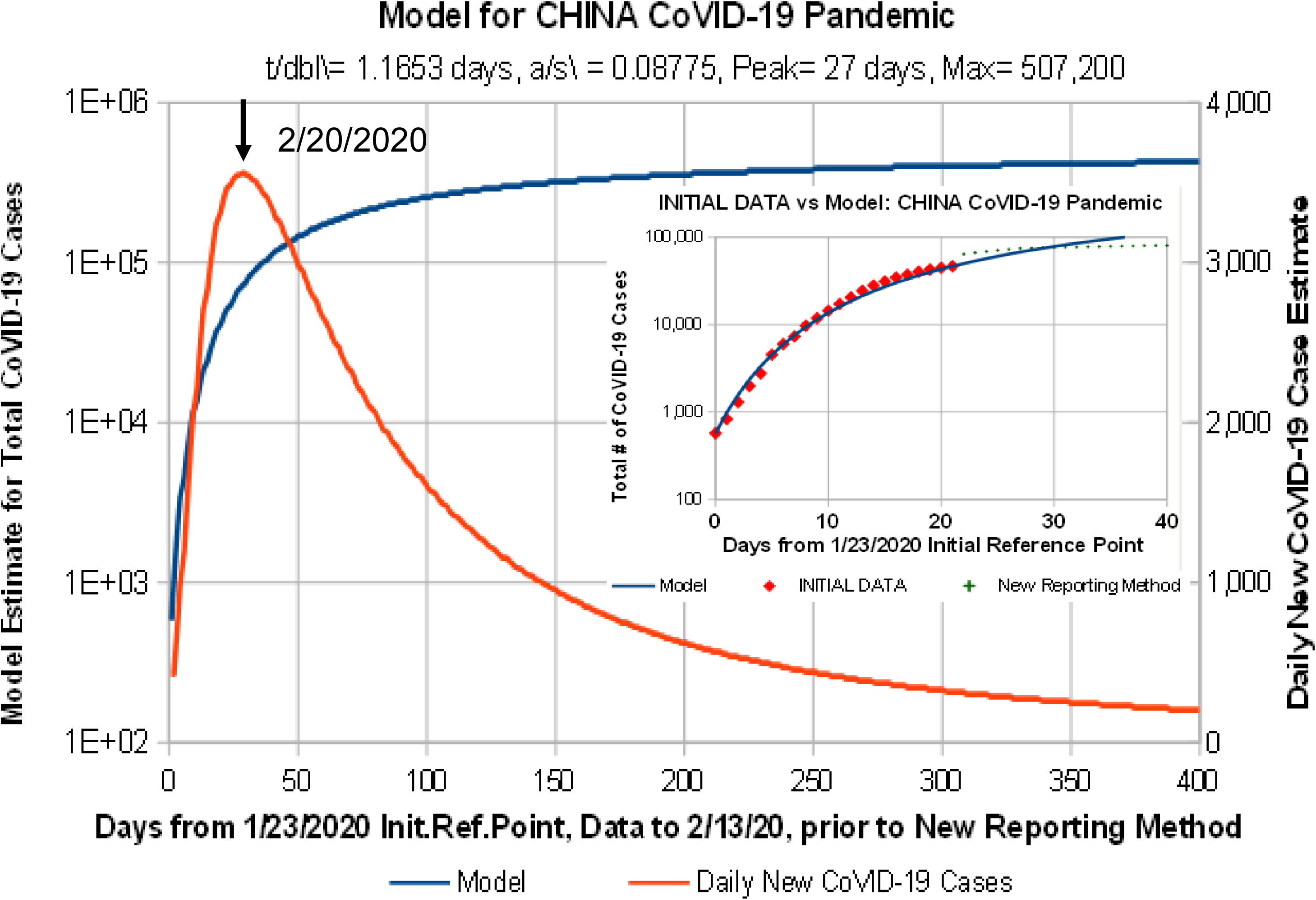
Predicted CHINA CoVID-19 results, using pre-*“New Reporting Method”* data. Draconian *Mitigation Measures* helped to contain pandemic to *Hubei Province* and *Wuhan*.

**Figure 11** shows model predictions for CoVID-19 evolution in South Korea, covering the period of their country’s early exposure and initial mitigation methods. Pre-pandemic *Mitigation Measures*, including extensive contact-tracing and large-scale CoVID-19 testing, were implemented. These projections show that those *Mitigation Measures*, as an alternative to China’s methods, also have impressively contained CoVID-19 spread.

**Figure 11:**
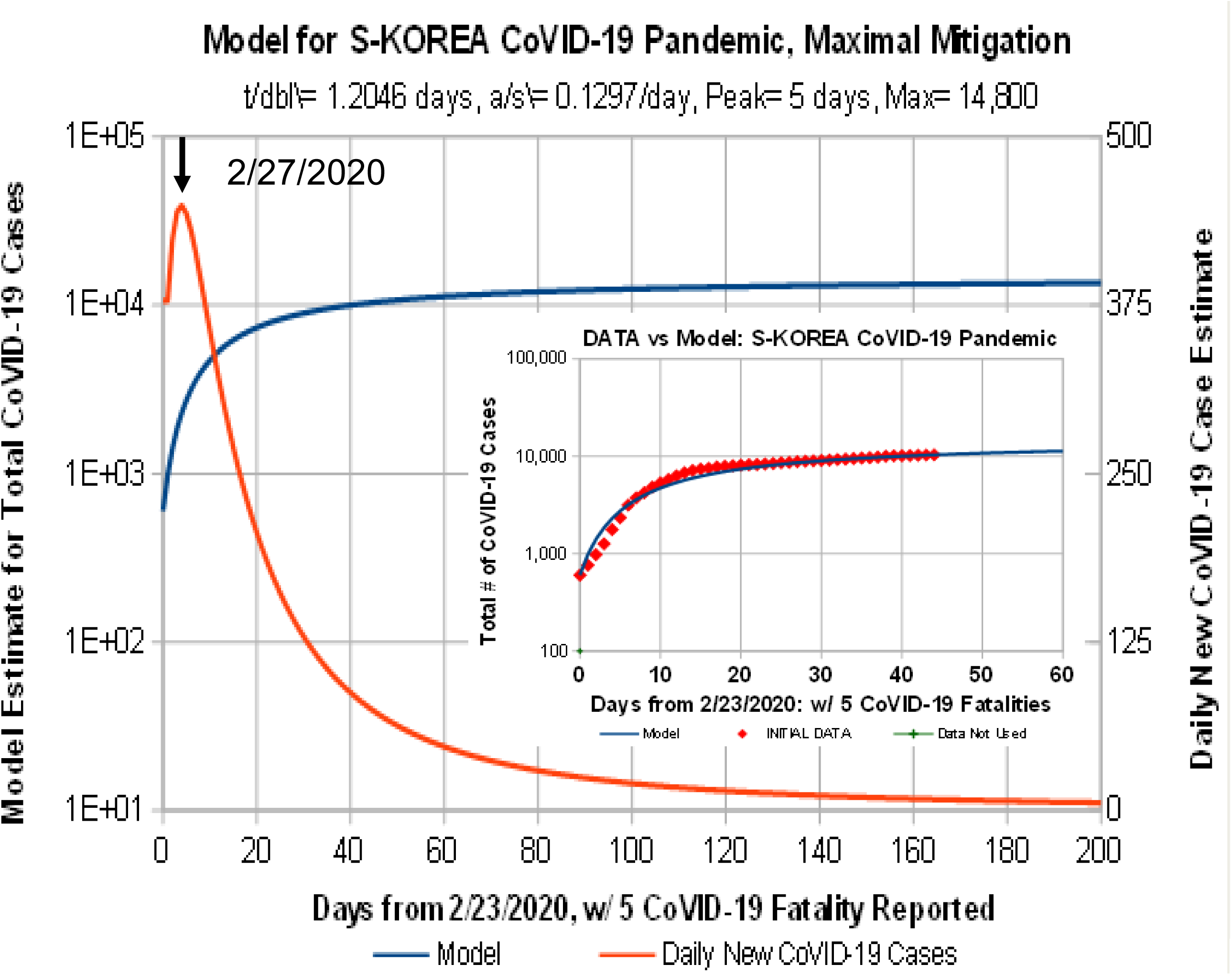
Predicted SOUTH KOREA CoVID-19 results. Pre-pandemic contact-tracing and large-scale CoVID-19 testing as *Mitigation Measures* have contained the pandemic.

**Figure 12** shows model predictions for CoVID-19 evolution in Italy. An initial *doubling-time* of 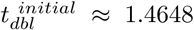 *days* and *SMP* estimate of *α_S_* ≈0.05282/day give a *pandemic peak* around 4/29/2020, with a predicted number of total cases at ^~^1, 764, 000, and a projected ^~^2.92% final infection rate.

**Figure 12:**
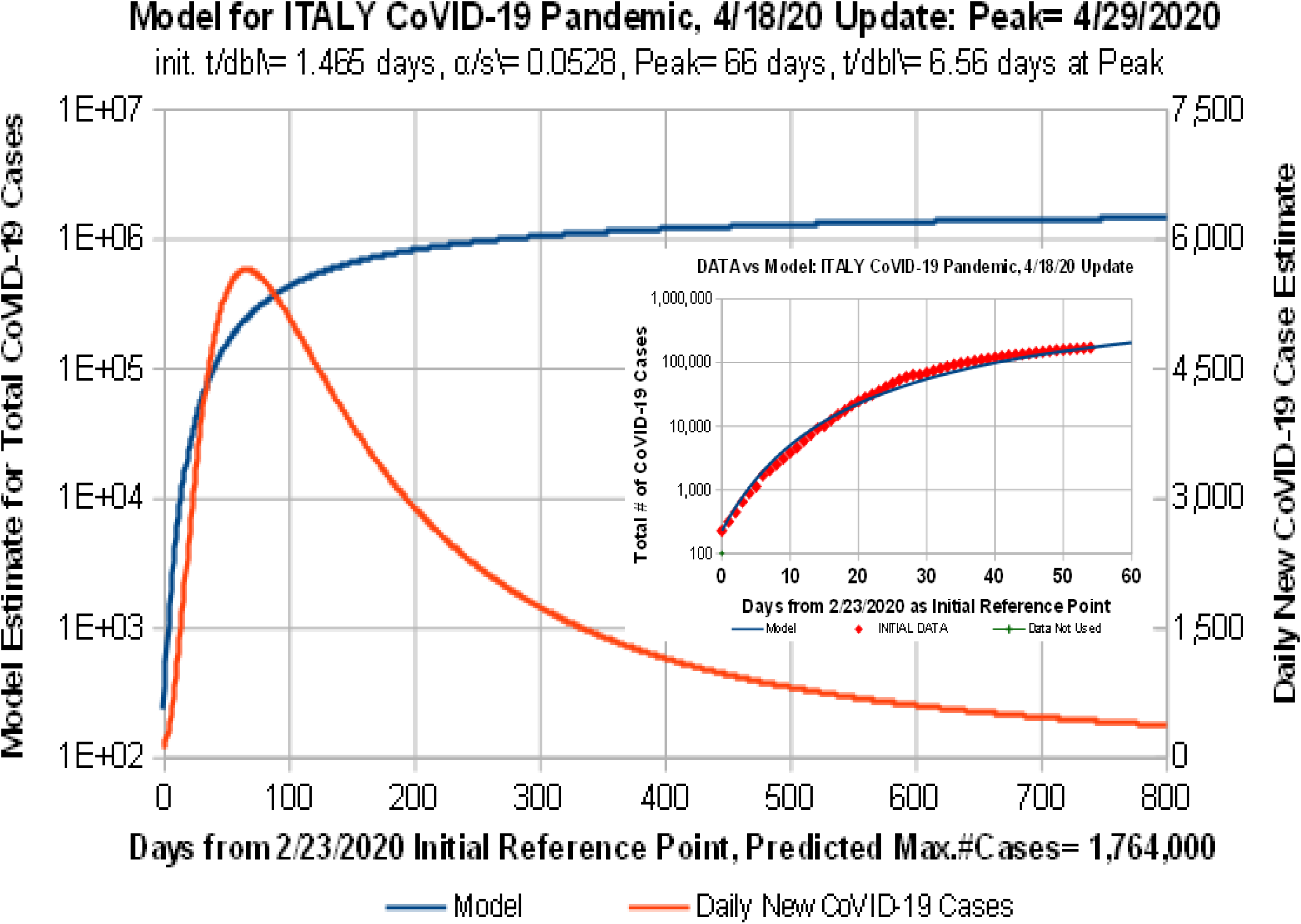
Predicted ITALY CoVID-19 results. Additional curvature in the actual CoVID-19 data vs Model makes these predictions a likely worst-case.

**Figure 13** shows model predictions for CoVID-19 evolution in Germany. The relatively high *SMP* estimate of *α_S_* ≈ 0.07614/day with an initial *doubling time* of 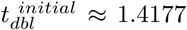 days combine to give a projected *pandemic peak* at around 4/08/2020, with a predicted total number of cases of ^~^700, 100, and a projected ^~^0.84% final infection rate. These values would make Germany one of the less impacted countries in Europe. They represent predicted final CoVID-19 infection rates that are significantly lower than the original 60% – 70% early worst-case estimates highlighted by *German Chancellor Angela Merkel*.

**Figure 13:**
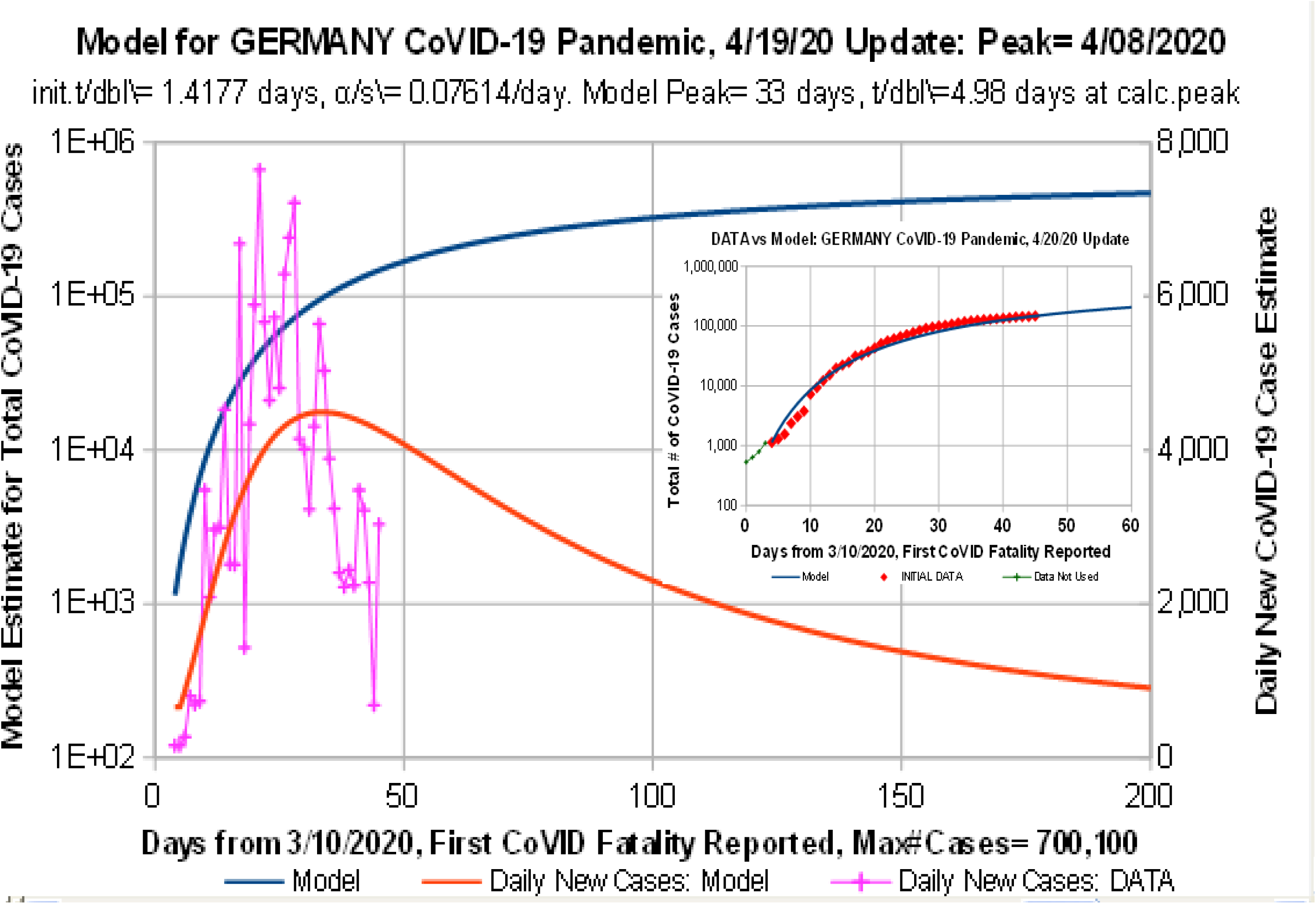
Predicted GERMANY CoVID-19 results. This model gives a more gradual function for the Daily New CoVID-19 cases, making these predictions a likely worst-case.

**Figure 14** shows model predictions for CoVID-19 evolution in Spain. An *SMP* estimate of *α_S_* ≈ 0.07058/*day*, which is comparable to Germany, and a smaller initial *doubling-time* of 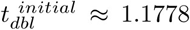 *days* combine to give more predicted CoVID-19 cases than Germany. The estimated *pandemic peak* is around 4/21/2020, with a predicted number of total cases at ^~^1, 526, 000, and a projected ^~^3.26% final infection rate.

**Figure 14:**
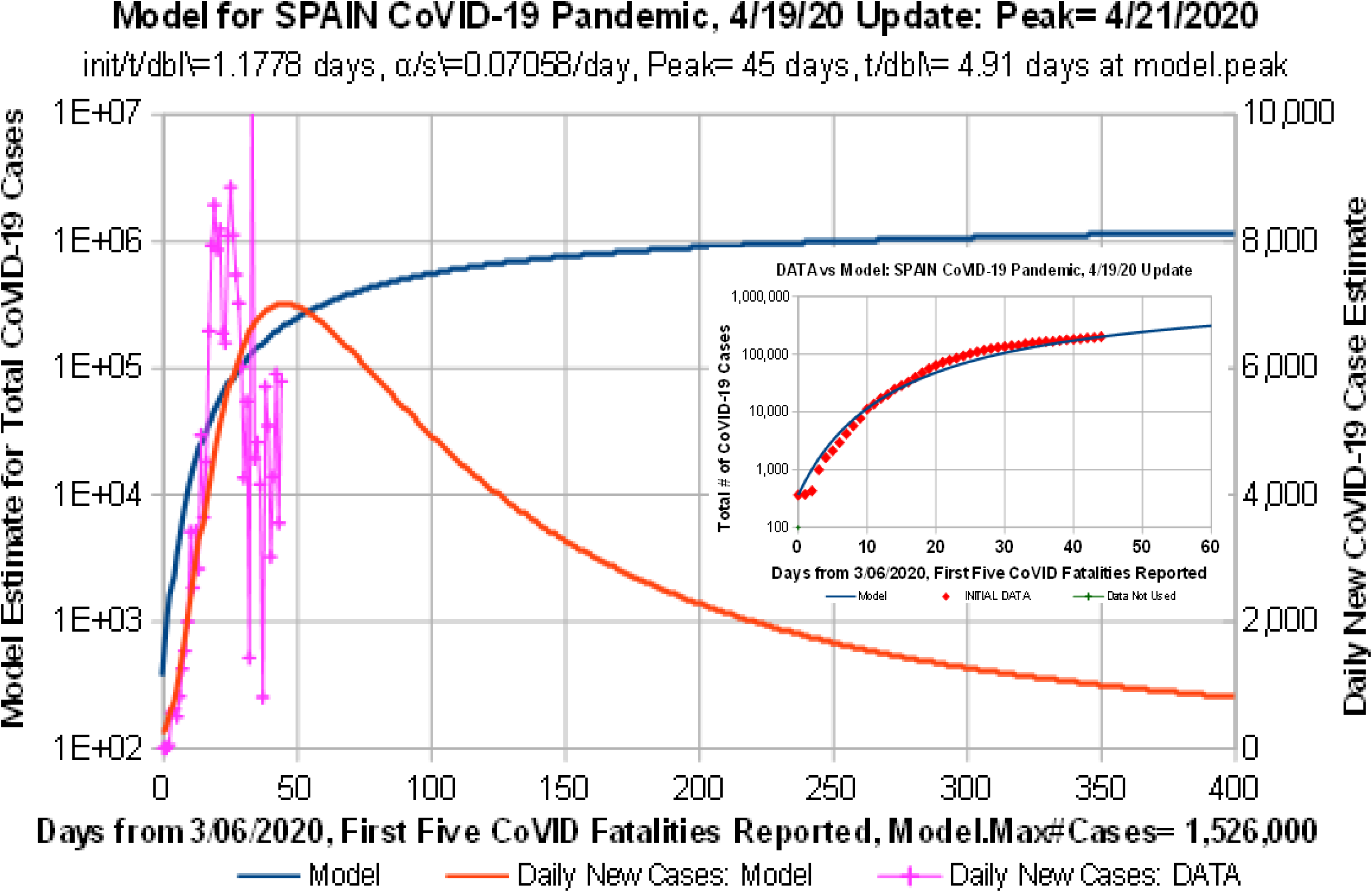
Predicted SPAIN CoVID-19 results. This model gives a more gradual function for Daily New CoVID-19 cases, making these predictions a likely worst-case.

**Figure 15** shows model predictions for CoVID-19 evolution in Ecuador. Reports of chaos in Ecuador have been alarming. Yet the present data show a significant and somewhat unexpected leveling off in the number of reported CoVID-19 cases. This result could mean that some as yet unknown *Mitigation Measures* may be operating. Alternatively, the data could mean that there is a dire CoVID-19 testing and reporting shortfall operating amidst the chaos.

**Figure 15:**
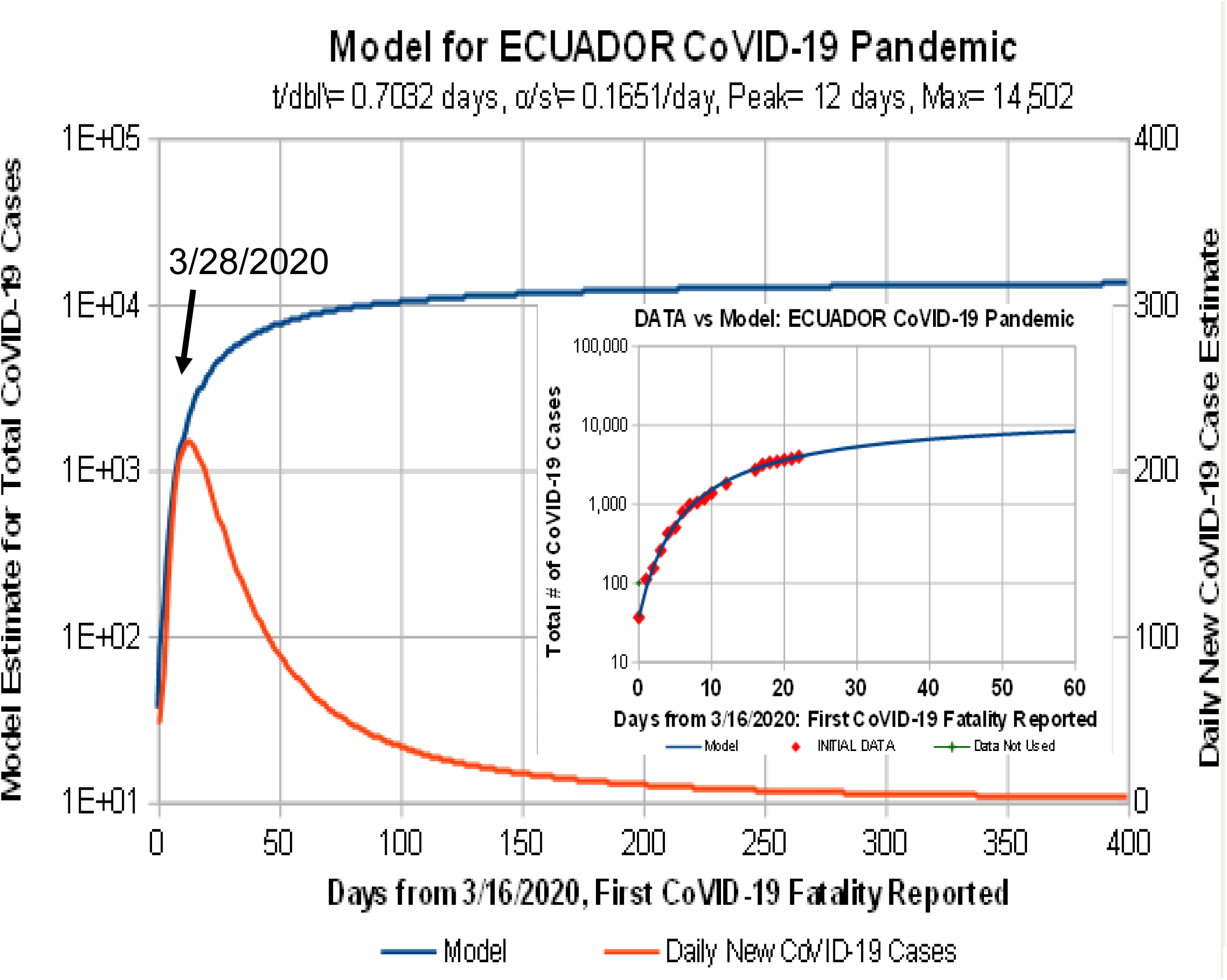
Predicted ECUADOR CoVID-19 results. Reports of chaos in Ecuador have been alarming. Poor CoVID-19 tracking and low testing may have skewed these results.

**Figure 16** shows model predictions for CoVID-19 evolution in India. These initial data show virtually no mitigation at present, having one of the lowest calculated *SMP* estimates of *α_S_* ≈ 0.0148/day, with an initial *doubling-time* of 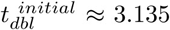 *days*. At this rate, nearly 17.38% of the population of India could eventually become infected. The estimated *pandemic peak* is around 5/30/2021, which would be 441 days after the first CoVID-19 fatality was reported, on 3/14/2020. Additional *Mitigation Measures*, further increasing the {*t_dbl_*, *α_S_* } values, as well as adding in additional modeling parameters may significantly reduce these projected number of CoVID-19 cases.

**Figure 16:**
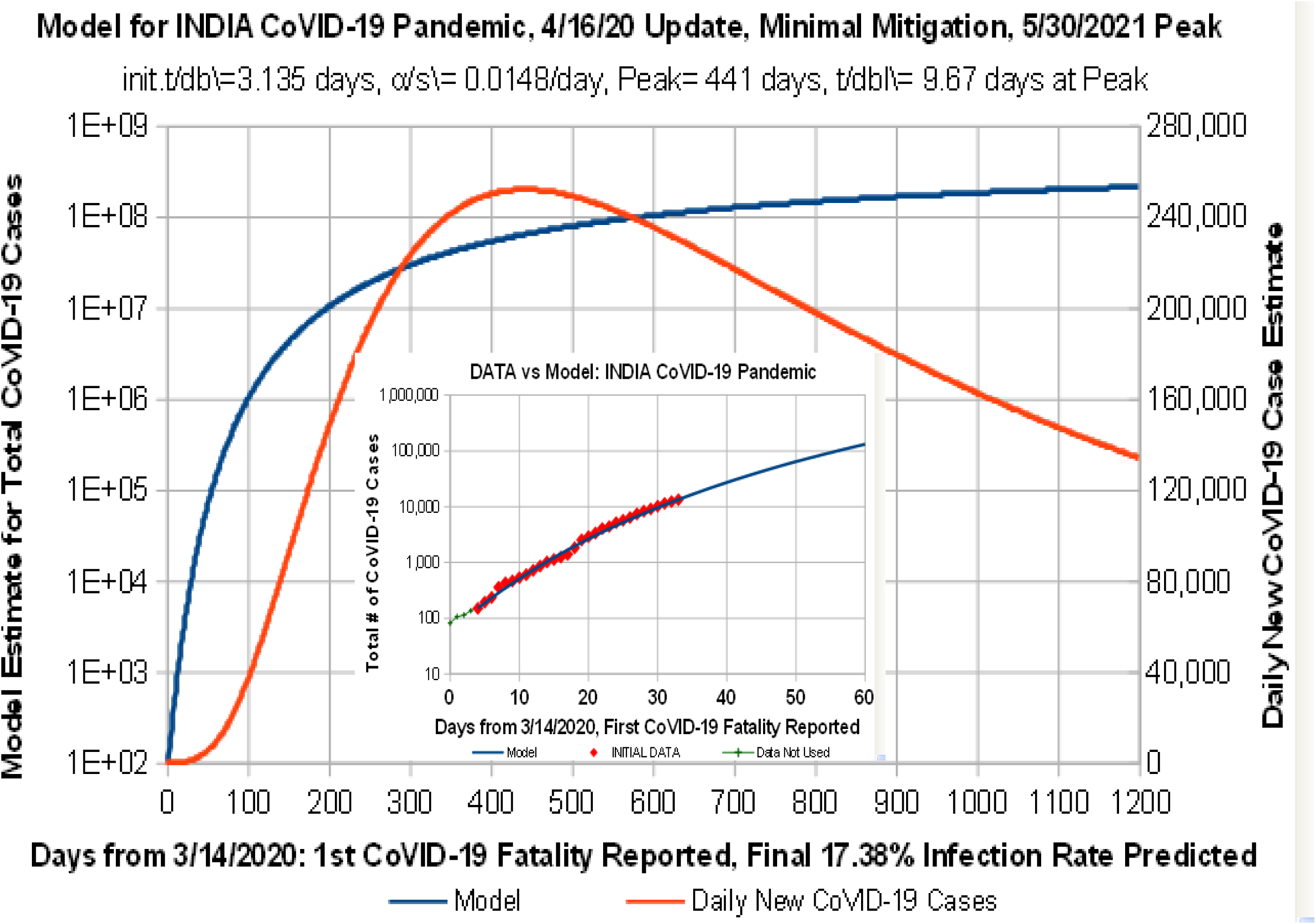
Predicted INDIA CoVID-19 results. Data shows only minimal mitigation at present. Further mitigations should help make these predictions a worst-case result.

## 7 Augmented Peak Shape Modeling

Using a new *Social Mitigation Parameter [SMP] α_S_*, as in Eq. [1.3a], successfully models pandemic shut-off, even in the *dilute pandemic* limit. However, as the **Figures 3-16** insets show, many of the data-vs-model comparisons have the data trending above the model near the final *t* = *t_F_* data point.

Since Eq. [1.3a] for *t_D_*(*t*) is linear, using an *Additional Modeling Parameter [AMP]* [*β*_S_ in a higher order polynomial, may fit the {*pdf*} shape better. A quadratic function for *t_D_*:

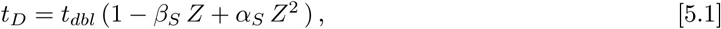

where *N*(*t*) / *N_o_* still approaches a constant at long times, as in Eq. [2.1a], then sets *Z*^2^ ≡ *t*, giving this extension of Eq. [1.3a]:

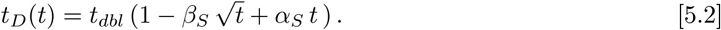

Values of [*β_S_* > 0 in Eq. [5.2] allow the predicted [*N*(*t*) / *N_o_*] values to rise above the [*β_S_* = 0 model predictions, and to have a smaller *doubling-time*, for the same {*t_dbl_*, *α_S_*}. However, the best fit {tdb, *α_S_*} values will also differ between the [*β_S_* > 0 and [*β* = 0 cases, so these changes are relative.

The new {*pdf*} function for Eq. [5.2] is:

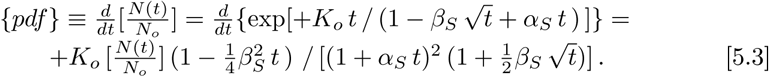

When {*pdf*} = 0 in Eq. [5.3], it estimates an end-point for the pandemic at:

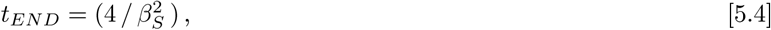

while predicting this maximum number of pandemic cases at *t_END_*:

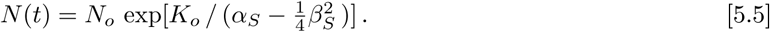

An an example, this augmented model was applied to CoVID-19 evolution in Italy. As shown in **Figure 17**, this Eq. [5.3] {*pdf*} function gives a better fit to the observed number of daily new CoVID-19 cases.

**Figure 17:**
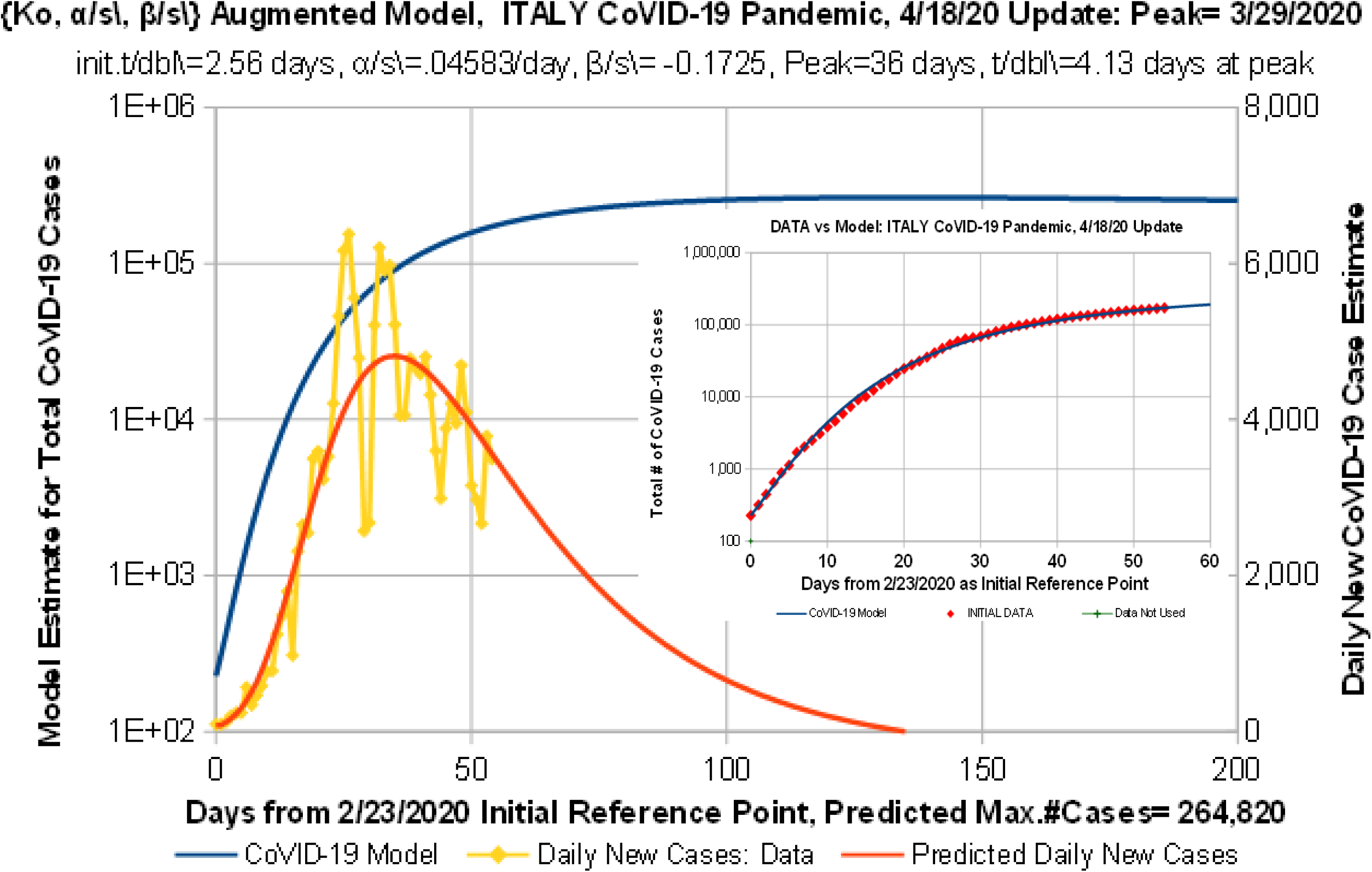
Predicted ITALY CoVID-19 results, using an augmented 2-parameter {*α_S_*, *β_S_*} *Social Mitigation* model. Total number of CoVID-19 cases is much less than **Figure 12**, but the model post-peak drop is much steeper, making this a likely best-case result.

The initial *doubling-time* of 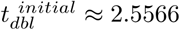*days*, along with estimates for the *Mitigation Measure* parameters of *α_S_* ≈ 0.04583/*day* and [*β_S_* ≈ −0.1725, in this augmented model, combine to significantly reduce the projected maximum number of CoVID-19 cases down to ^~^264, 820, which is an ^~^7*X* less compared to using *α_S_* alone, as in **Figure 12**. This augmented model sets an estimated *pandemic peak* at 3/29/2020, with a projected pandemic end-point around 7/7/2020, which is also significantly more optimistic.

The true CoVID-19 pandemic progress is likely to be in between **Figure 12** as a worst-case, and **Figure 17** as a best-case projection. The geometric mean of the **Figure 12** and **Figure 17** results set an average of ^~^683, 500 cases for Italy at the CoVID-19 pandemic end. These bounds also highlight the amount of uncertainty that is intrinsic to these empirically based methods.

## 8 Summary and Conclusions

The standard exponential for modeling pandemics starts with an *N_o_* known number of initial cases at some reference time *t* = 0. Epidemiologists work to determine a pandemic growth factor *K_D_*, which sets the *doubling-time t_D_* for the number of pandemic cases.

The CoVID-19 disease, caused by the SARS-CoV-2 coronavirus pathogen, initially showed both regional and global exponential growth. It resulted in a *doubling-time* of *t_D_* _≈_ 2.02 *days* for the US, as highlighted in **Figure 1**.

An exponential growth normally only halts when it runs out of materials. In epidemiology that point often occurs when there are virtually no more uninfected people left, which we call a *saturated pandemic*. The exponential growth function is only applicable when infection rates are much lower than saturation, which we call a *dilute pandemic*.

A modification to exponential growth is developed here, which allows ratio of the number of pandemic cases, *N*(*t*), compared to its *N_o_* initial value at *t* = 0:

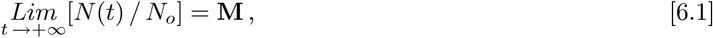

to approach a final constant, denoted **M**, while still being in a *dilute pandemic* condition. This result is attributed to the inclusion of society-wide *Mitigation Measures* to stop pandemic growth, before the value of **M** reaches the whole population value.

Society-wide *Mitigation Measures* aim to progressively lengthen the *t_D_ doubling time*, essentially making *t_D_* (*t*). Most analyses presented here used a linear function of time as a simplest non-constant model for *t_D_* (*t*):

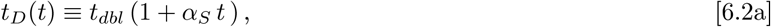

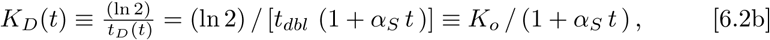

where *t_D_*(*t* = 0) = *t_dbl_*, and *K_D_*(*t* = 0) = *K_o_*. Here, as is a new *Social Mitigation Parameter (SMP)*, to quantify societal *Mitigation Measures*. This Eq. [6.2a] extension of a pure exponential growth gives:

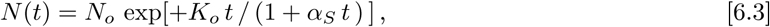

as an empirical equation for modeling CoVID-19 spread. Since both *K_o_* and *α_S_* in Eq. [1.4] have the same units, their ratio is a dimensionless number. The long-term limit of Eq. [6.3] gives:

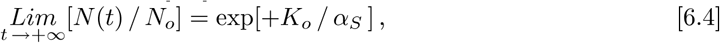

setting a final value for the Eq. [6.1] constant **M**, allowing these predictions to be applicable to the *dilute pandemic* limit. The CoVID-19 number of estimated cases per day is given by:

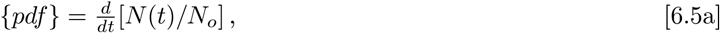

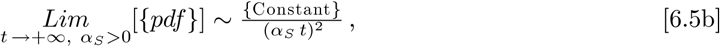

which combines an initial exponential rise with “*long tail*” at large times. In this model, new CoVID-19 cases can continue to arise for a long time, even with significant *Mitigation Measures* in place.

Analysis of available CoVID-19 data using this model shows that it can match observed data fairly well, both from various US states [**Figures 3-8**], as well as for different global countries [**Figures 9-17**]. However, using a single parameter to encompass all societal *Mitigation Measures* often gives a slightly larger slope on a *log-plot*, compared to the latest measured data values, which makes this model a likely worst-case estimate.

A second data-fitting parameter *β_S_* was also used in an augmented model, to better fit the {*pdf*} data:

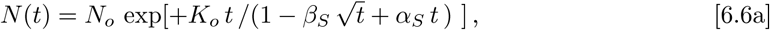

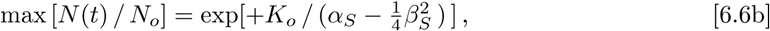

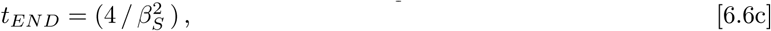

where *t_END_* becomes an estimated pandemic end-point, where zero new CoVID-19 cases per day could occur.

As a representative example, this augmented model was applied to the CoVID-19 data from Italy [**Figure 17**]. Those results show that this augmented model allows a better fit to the observed number of new daily CoVID-19 cases, but the absence of a CoVID-19 tail in its {*pdf*} function makes this {*K_o_*, *α_S_*, *β_S_*} augmented model a likely best-case result, with the original {*K_o_*, *α_S_*} model being a likely worst-case estimate.

This class of CoVID-19 pandemic models all enable pandemic shut-off even in the *dilute pandemic* limit, with only a small fraction of the total population being infected. These models also provide estimates for: (a) the maximum number of cases near pandemic shutoff, (b) the size and shape of the *pandemic peak* [*dN*(*t*) / *dt*], and (c) pandemic peak timing [*t_P_*]. These models and analyses may help enhance planning and preparation to maximize resource use, potentially increasing individual and collective CoVID-19 pandemic survival rates.

## Data Availability

CC BY: All data and analyses are publicly available

